# Alpha-synuclein distribution and seeding activity in rectal biopsies in Parkinson’s disease

**DOI:** 10.1101/2024.03.29.24304902

**Authors:** Annika Kluge, Carmen Kintrup, Kristina Kulcsarova, Katja Schröder, Julius Welzel, Sebastian Heinzel, Thilo Wedel, Martina Böttner, Ralph Lucius, Sarah Kim Bonkat, Manuela Pendziwiat, Stephan Schoch, Mark Ellrichmann, Daniela Berg, Eva Schaeffer, François Cossais

**Affiliations:** Department of Neurology, Kiel University, University Medical Center Schleswig-Holstein, Kiel, Germany; Institute of Anatomy, Kiel University, Kiel, Germany; Department of Neurology, P.J. Safarik University and L. Pasteur University Hospital, Kosice, Slovak Republic; Department of Clinical Neurosciences, University Scientific Park MEDIPARK, P. J. Safarik University, Kosice, Slovak Republic; Institute of Medical Informatics and Statistics, Kiel University, Kiel, Germany; Institute of Clinical Molecular Biology, Kiel University, Kiel, Germany; Interdisciplinary Endoscopy, Medical Department I, Kiel University, University Medical Center Schleswig-Holstein, Kiel, Germany

**Author notes:** These authors contributed equally to this work (Eva Schaeffer, François Cossais).

## Abstract

**Background:** Parkinson’s disease (PD) is characterized by the accumulation of alpha-synuclein (aSyn) pathology, not only in the brain but also in the gastrointestinal (GI) tract. This study investigates the use of unique aSyn antibodies and an aSyn seed amplification assay (SAA) for detecting pathological aSyn in rectal biopsy samples from PD patients and healthy individuals. These samples were preserved using formalin-fixed paraffin-embedded (FFPE) methods.

**Materials and Methods:** The study analyzed the seeding capacity of FFPE submucosal rectal biopsies from 24 PD patients and 20 healthy controls using an aSyn-SAA. The distribution of aSyn was examined using immunohistochemistry with antibodies targeting specific conformations and phosphorylated forms of aSyn at S129 and Y39.

**Results:** Pathological forms of aSyn were found in all FFPE biopsies from PD patients, as confirmed by SAA, and these were linked to the severity of motor symptoms (MDS-UPDRS-III). However, the immunoreactive patterns of conformation-specific or phosphorylated aSyn in rectal biopsies did not show notable differences between PD patients and healthy subjects.

**Conclusion:** Pathological aSyn strains are detectable in FFPE rectal biopsies from PD patients with high accuracy using aSyn-SAA. However, the utility of immunohistochemical detection with current antibodies for identifying pathological aSyn forms appears limited. The findings advocate the use of aSyn-SAA as a diagnostic tool for PD, contributing to a deeper understanding of the gut-brain connection in the disease.

## Introduction

The majority of patients with Parkinson’s disease (PD) present with gastrointestinal (GI) disorders, including constipation or gastroparesis (1). Pathological aggregation of alpha-synuclein (aSyn) in Lewy bodies is the histological hallmark of PD. Corresponding to its clinical presentation, the expression of aSyn is not limited to the central nervous system (CNS), but is also observed in the periphery including the enteric nervous system (ENS) (2,3). In fact, aSyn deposits in association with low-grade inflammation have been found in the intestinal tissue of PD patients (4–6). A careful analysis of pathological aSyn deposits in peripheral organs and the brain led Braak and co-authors to hypothesize that PD may spread from the periphery to the CNS (7). Based on this assumption, significant efforts have been made to detect misfolded forms of aSyn in GI tissues *ex vivo* in order to comprehend underlying pathophysiological mechanisms of the gut-brain axis and evaluate enteric aSyn expression as a potential diagnostic biomarker for PD (8).

Despite conflicting results, there is a global consensus about the fact that misfolded aSyn may be present along the GI tract in most PD patients (5,9,10). Nonetheless, the characterization of pathological aSyn in GI tissue has been challenging; the methods used for this purpose have been diverse (2,3,8,10–12). Histological approaches to detect aSyn are hindered by the complex diversity of aSyn conformation forms, moreover, little is known about the aSyn conformational state along the gut-brain axis under physiological conditions or in PD. So far, histological approaches targeting GI aSyn have been aimed at detecting phosphorylated forms of aSyn at position S129 (P-S129), as this post-translational modification was shown to be present in high levels in Lewy bodies (13). Additionally, the recent development of conformation-specific aSyn antibodies offers a novel opportunity to detect misfolded forms of aSyn at the histological level (14). Immunoreactive patterns observed with a conformation-specific aSyn antibody, clone 5G4, in the duodenum and the colon appears to distinguish between PD and control tissues with high specificity and sensitivity (15,16). However, the specificity of aSyn phosphorylation- and conformation-specific antibodies has been questioned and their validity in immunohistochemical staining of GI tissue has not been fully evaluated yet (17).

The recent development of aSyn seed amplification assays (SAA), encompassing real-time quaking-induced conversion (RT-QuIC) and the protein misfolding cyclic amplification assay (PMCA), which are based on the *in vitro* amplification of tissue-derived misfolded aSyn seeds (18,19), has emerged as an additional alternative method for the detection of aSyn pathology in intestinal tissues (20–22).

In the present report we evaluated the potential of SAA in comparison to phosphorylation-specific antibodies and conformation-specific antibodies in demonstrating misfolded aSyn in formalin-fixed and paraffin-embedded (FFPE) rectal biopsies of PD patients and controls.

## Material and methods

### Patients

Twenty-four patients with PD who were diagnosed according to the Movement Disorder Society (MDS) Clinical Diagnostic Criteria for Parkinson’s Disease (23) and 20 healthy individuals (controls) were included in the study (Table 1). All individuals underwent colonoscopy for colorectal cancer screening at the Interdisciplinary Endoscopy, Department of internal medicine, University Hospital Schleswig-Holstein, Campus Kiel.

**Table 1:** Clinical data and results.

|  | Controls (n=20) | PD (n=24) | Sig. PD vs controls (p) |
| --- | --- | --- | --- |
| **Sex m/f** | 9/11 | 20/4 | 0.008 <sup>*b</sup> |
| **Age** years, mean ± SD | 66.2 ± 8.1 | 65.3 ± 8.3 | 0.417 <sup>a</sup> |
| **Disease duration** years, mean ± SD | - | 7.0 ± 6.0 | - |
| **LEDD** mean ± SD | - | 410.2 ± 367.1 | - |
| **MDS-UPDRS part III**<br>total score, mean ± SD, (range) | 0.6 ± 1.3<br>(0-5) | 25.2 ± 13.5<br>(2-54) | < 0.001 <sup>*a</sup> |
| **Wexner's constipation score**<br>total score, mean ± SD, (range) | 3.2 ± 3.7<br>(0-10) | 4.5 ± 4.5<br>(0-15) | 0.154 <sup>a</sup> |
| **SAA** |  |  |  |
| - Positivity (n, %) | 0/20 (0%) | 24/24 (100%) | < 0.001 <sup>*b</sup> |
| - F<sub>max</sub> (AU) | 2.0 (1.2-3.8) | 75.1 (53.6-99.3) | < 0.001 <sup>*a</sup> |
| - T<sub>50</sub> (h) | 20.3 (0.0-48.0) | 31.4 (29.4-35.6) | 0.262 <sup>a</sup> |
| - AUC (AU) | 77.7 (41.6-168.8) | 1350.9 (955.8-1594.5) | < 0.001 <sup>*a</sup> |
| - Lag phase (h) | n.a. | 21.2 (10.0-24.0) | n.a. |
| **IHC number (%)** |  |  |  |
| - **syn211** |  |  |  |
| - neuronal | 5 (25) | 8 (33) | 0.546 |
| - neurites | 4 (20) | 6 (25) | 0.974 |
| - ganglia | 2 (10) | 6 (25) | 0.117 |
| - Non-neuronal (mucosal) | 4 (20) | 4 (17) | 0.775 |
| - **N-term aSyn** |  |  |  |
| - neuronal | 5 (25) | 5 (21) | 0.743 |
| - neurites | 3 (15) | 4 (17) | 0.880 |
| - ganglia | 2 (10) | 2 (8) | 0.848 |
| - Non-neuronal (mucosal) | 20 (100) | 24 (100) | n.a. |
| - **MJF-R14** |  |  |  |
| - neuronal | 20 (100) | 20 (83) | 0.056 |
| - neurites | 20 (100) | 20 (83) | 0.056 |
| - ganglia | 7 (35) | 4 (17) | 0.162 |
| - Non-neuronal (mucosal) | 20 (100) | 24 (100) | n.a. |
| - **5G4** |  |  |  |
| - neuronal | 0 (0) | 1 (4) | 0.356 |
| - neurites | 0 (0) | 1 (4) | 0.356 |
| - ganglia | 0 (0) | 0 (0) | n.a. |
| - Non-neuronal (mucosal) | 15 (75) | 12 (50) | **0.039** |
| - **P-S129 aSyn (MJF-R13)** |  |  |  |
| - neuronal | 16 (80) | 20 (83) | 0.498 |
| - neurites | 16 (80) | 20 (83) | 0.498 |
| - ganglia | 8 (40) | 3 (12) | **0.036** |
| - Non-neuronal (mucosal) | 19 (95) | 24 (100) | 0.268 |
| - **P-S129 aSyn (pSyn#64)** |  |  |  |
| - neuronal | 7 (35) | 18 (75) | **0.001** |
| - neurites | 7 (35) | 18 (75) | **0.001** |
| - ganglia | 4 (20) | 14 (58) | **0.010** |
| - Non-neuronal (mucosal) | 18 (90) | 24 (100) | 0.113 |
| - **P-Y39 aSyn** |  |  |  |
| - neuronal | 3 (15) | 1 (4) | 0.213 |
| - neurites | 0 (0) | 0 (0) | n.a. |
| - ganglia | 3 (15) | 1 (4) | 0.213 |
| - Non-neuronal (mucosal) | 18 (90) | 19 (79) | 0.328 |

AU: arbitrary units; PD: Parkinson's disease; h: hours; LEDD: L-Dopa equivalent daily dose; SAA: alpha-synuclein seed amplification assay; Sig.: statistical significance, <sup>a</sup> Mann-Whitney U test, <sup>b</sup> Chi-square test. \* statistically significant result. n.a.: not applicable

The MDS Unified Parkinson’s Disease Rating Scale (MDS UPDRS-III) and Wexner’s constipation score were obtained for both groups. The levodopa-equivalent daily dose (LEDD) was determined for PD patients. The study was approved by the local ethics committee of the faculty of medicine, Christian-Albrechts University of Kiel, Germany (D452/19).

Additionally, a substantia nigra specimen was obtained from two body donors with PD and one control individual without known neurological disorders, as detailed in the supplementary methods.

### Retrieval and Processing of rectal biopsies

From each individual, we obtained two deep submucosal biopsies from the rectum during screening colonoscopy (Jumbo Radial Jaw 3.2 mm biopsy forceps; Boston Scientific, Malborough, MA, USA). The samples were fixed in 4% paraformaldehyde in phosphate-buffered saline (PBS) prior to paraffin embedding. Paraffin-embedded tissue blocks were cut in sections of 6 µm thickness, transferred onto glass slides, and stored at room temperature until further processing. Immunohistochemical analyses and aSyn-SAA were performed for all 24 PD patient samples and 20 healthy control samples as detailed below.

### Retrieval and Processing of Substantia nigra specimens

Substantia nigra tissue was obtained post-mortem from two body donors with diagnosed PD, as well as from one control individual without known neurological manifestation (two females, one male respectively, age range: 88-93 years), who were recruited from the body donation program of the Institute of Anatomy, Kiel University, Germany. All samples were fixed in 4% paraformaldehyde in PBS until they were embedded in paraffin. The paraffin embedded tissue-blocks were then cut into 6 µm thick sections that were transferred onto glass slides and stored at room temperature until further processing.

### Alpha-synuclein seed amplification assay

The seed amplification assay (SAA) was performed on FFPE material obtained from histological slides of the 24 PD patients and 20 controls. The slides were soaked in Tris buffer (150 mM KCl, 50 mM Tris-HCl, pH 7.5) for 10 minutes and the paraffin sections were removed from the slides with a scalpel. For each patient or control, 30 histological sections (6 µm thickness) were pooled and collected in 150 µl Tris buffer. The tissue was dissolved by incubation in Tris buffer for at least 72 hours in a plate shaker at 650 rpm and 37 °C, with repeated centrifugation and mixing steps using a vortex mixer. After several rounds of centrifugation at 15,000 x g and washing with Tris buffer, the homogenized lysates were used for the SAA. The homogenized FFPE material (10 mg) was incubated with 500 µg of recombinant monomeric aSyn (#S-1001-2, rPeptide, Watkinsville, USA) in 100 µl PBS buffer in a dark 96-well plate (#437111, Thermo Fisher Scientific). The plates were then covered with silicone lids (Thermo Fisher Scientific, #AB0566) and PARAFILM M sealing film (Bemis, USA), incubated at 36.5 °C, agitated at 1,000 rpm using a plate shaker (MTS 4, IKA), and placed in an incubator. Thioflavine T (ThT) at a final concentration of 0.01 mM was freshly prepared before each measurement using 1 mM stock solution. For each measurement, 1 µl of ThT was manually added to each well so the total ThT content increased with each measurement. The ThT fluorescence signal intensity was measured at indicated time points which were determined according to the curve of the positive control, and stopped when the ThT fluorescence plateau was reached. The ThT fluorescence signal intensity was monitored at the excitation wavelength of 410 nm and the emission wavelength of 475 nm using a microplate reader (Infinite 200 PRO, Tecan) and quantified in arbitrary units (A.U.). The plates were replaced on the shaker and incubated at 36.5 °C during the time between measurements. As a positive control, 3.4 ng of pre-formed aSyn fibrils were incubated with 500 µg aSyn monomers. As negative controls we used 500 µg aSyn monomers without further seeding material, as well as 3.4 ng of pre-formed fibrils without incubation with monomeric aSyn. The average of ThT signal raw data during the first 10 hours of recording of all control samples plus 5 standard deviations was calculated to determine the threshold for positive seeding. ThT values per patient as inputs were used to determine the following measures after linear interpolation of data: peak of the ThT fluorescence response (F_max_), time to reach 50 % of F_max_ (T_50_), area under the curve (AUC), and the lag phase (time taken to reach the threshold). Analyses were performed with custom scripts using a Python 3.10.11 environment, including numpy v1.24.3 for numerical computations, pandas v2.0.2 for data manipulation and analysis, and scipy v1.10.1 for scientific computations.

### Immunohistochemical staining

Immunohistochemical staining was performed in accordance with a standard protocol (4). Briefly, sections were pretreated with xylol for deparaffinization, alcohol with descending concentrations for rehydration, and citrate buffer (pH=6.0) to unmask antigens. For staining with clone 5G4 (14), the sections were incubated for 1 minute in 98% formic acid (Sigma). Sections were rinsed with distilled water and PBS buffer (pH=7.4), and incubated overnight with the primary antibodies (Table S1) in antibody diluent (Thermo Fisher Scientific, Carlsbad, USA). After washing with PBS, the sections were incubated for one hour with the secondary antibodies. For immunohistochemical investigation we used the BrightVision 1 step detection system anti-mouse HRP and anti-rabbit HRP (ImmunoLogic). Immpact DAB EqV was used as substrate for HRP (Vector Laboratories). Nuclei were counterstained with hematoxylin. Images were acquired using a Leica Aperio CS2 slide scanner or a Keyence BZ-x800e inverted microscope. Blank controls were performed for all tissue types by omitting primary antibodies. Full-thickness colon specimen and substantia nigra specimen processing and staining are detailed in the supplementary methods. Immunoreactive signal (IR) and tissue area was quantified using Aperio ImageScope software (v12.4.6.5003, Leica) running the “positive pixel count 2004-08-11” algorithm. Mean IR-positive area of five different mucosal fields for each biopsy was measured. Results are represented as percentage of the total measured area.

### Statistical analyses

Prism GraphPad version 9.0 and SPSS version 29 were used for statistical analysis. Statistical differences between two groups were evaluated by the t-test and for three or more groups using ANOVA followed by Tukey’s post-hoc tests. For SAA, differences between groups were calculated with the Mann-Whitney U test. Differences between PD and control groups for categorical variables were assessed with the Chi-square test. Test results with a p-value < 0.05 were considered statistically significant. Spearman’s rank correlation (two-sided) was applied for correlations in all PD patients, and while primary analyses aimed at correlations between F_max_ and AUC with PD motor symptoms severity (MDS-UPDRS-III) additional correlations were considered exploratory.

## Results

### Clinical data

The healthy control and PD group were comparable regarding age (mean age controls: 66, SD 8 years; PD: 65, SD 8 years; p = 0.42), while the PD group comprised more men (male sex controls: 45%, PD: 83%; p = 0.008). No group difference was seen for the Wexner’s constipation score.

### aSyn seeding activity of FFPE rectal biopsies from PD patients and healthy controls

The seeding activity of aSyn in FFPE rectal biopsies of PD patients was first assessed using aSyn-SAA (Fig. 1). All of the 24 PD patients and none of the healthy controls had a positive SAA (sensitivity 100%). F_max_ and AUC ThT values were significantly increased in PD patients compared to healthy controls (Table 1). No significant difference was seen for T_50_. In PD patients, F_max_ and AUC correlated significantly with MDS-UPDRS-III sum scores (F_max_: r = 0.463, p = 0.030; AUC: r = 0.525, p = 0.012; Fig. S2 and S3). F_max_ also showed a significant correlation with disease duration (r=0.421, p = 0.046), while no correlation with Wexner score or LEDD was observed. Clinical parameters showed no correlation with T_50_ or lag phase (Fig. S2).

**Figure 1:**
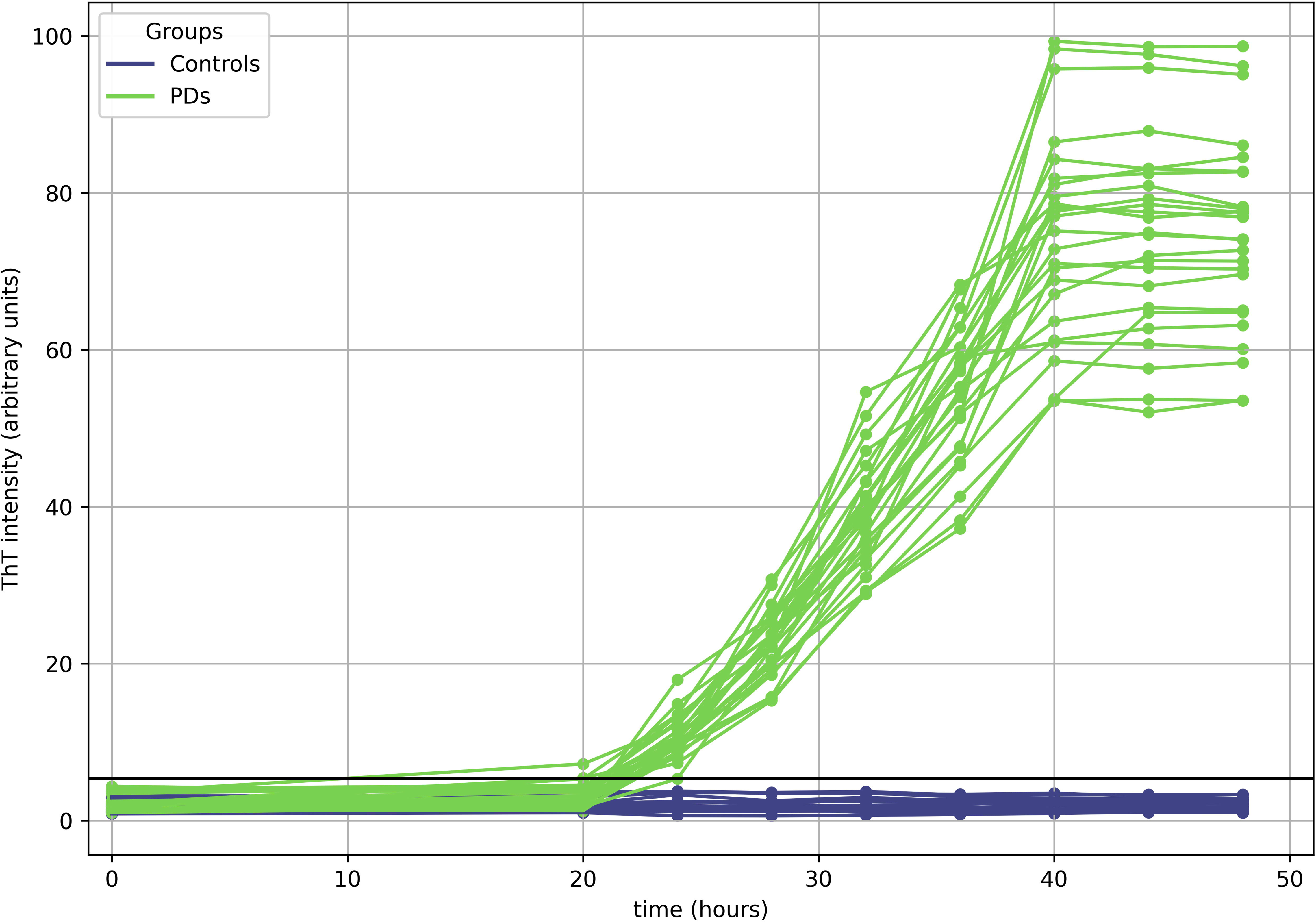
aSyn seeding activity of FFPE rectal biopsies of PD patients and control individuals. aSyn seed amplification assay was performed on FFPE rectal biopsies of PD patients (PD, green lines) and healthy subjects (Control, blue lines). ThT (Thioflavine T) signal intensity was measured over time. Threshold is indicated by a black line.

### Distribution pattern of aSyn in FFPE rectal biopsies of PD patients and healthy controls

In order to determine the presence of putative pathological aSyn deposits, we performed a characterization of aSyn distribution in intestinal tissues using pan-aSyn antibodies mapped to the C-terminal (syn211) or N-terminal part (N-term aSyn) of the aSyn protein respectively (Fig. S1). Immunoreactive patterns of all employed antibodies were first evaluated in the substantia nigra of two PD patients and one control (Fig. S4). Both the syn211 and N-term-aSyn antibodies, clearly revealed Lewy bodies (LB) and Lewy neurites (LN) in PD substantia nigra but not in control tissues, with the N-term antibody displaying strongest background staining (Fig. S4).

The distribution pattern of aSyn was then evaluated by immunohistochemistry in FFPE deep submucosal rectal biopsies of PD patients and healthy control individuals, which contained mucosal tissue as well as a part of the muscularis mucosae and submucosal connective tissue including submucosal ganglia. The presence of nerve fibers within the biopsies was first confirmed by staining for beta-III tubulin (TUBB3, Fig. S5). Immunoreactivity for aSyn was localized in enteric neurites and ganglia in about 25% controls and PD specimens, using either syn211 or N-term-aSyn antibodies (Fig. 2 a-b, Table 1). Immunoreactivity was also observed in non-neuronal cells within the mucosa in almost all specimen using the N-term-aSyn antibody (Table 1). The positive area covered after staining with syn211 or N-term-aSyn antibodies did not differ significantly between PD and control biopsies (Fig. 2 c-d).

**Figure 2:**
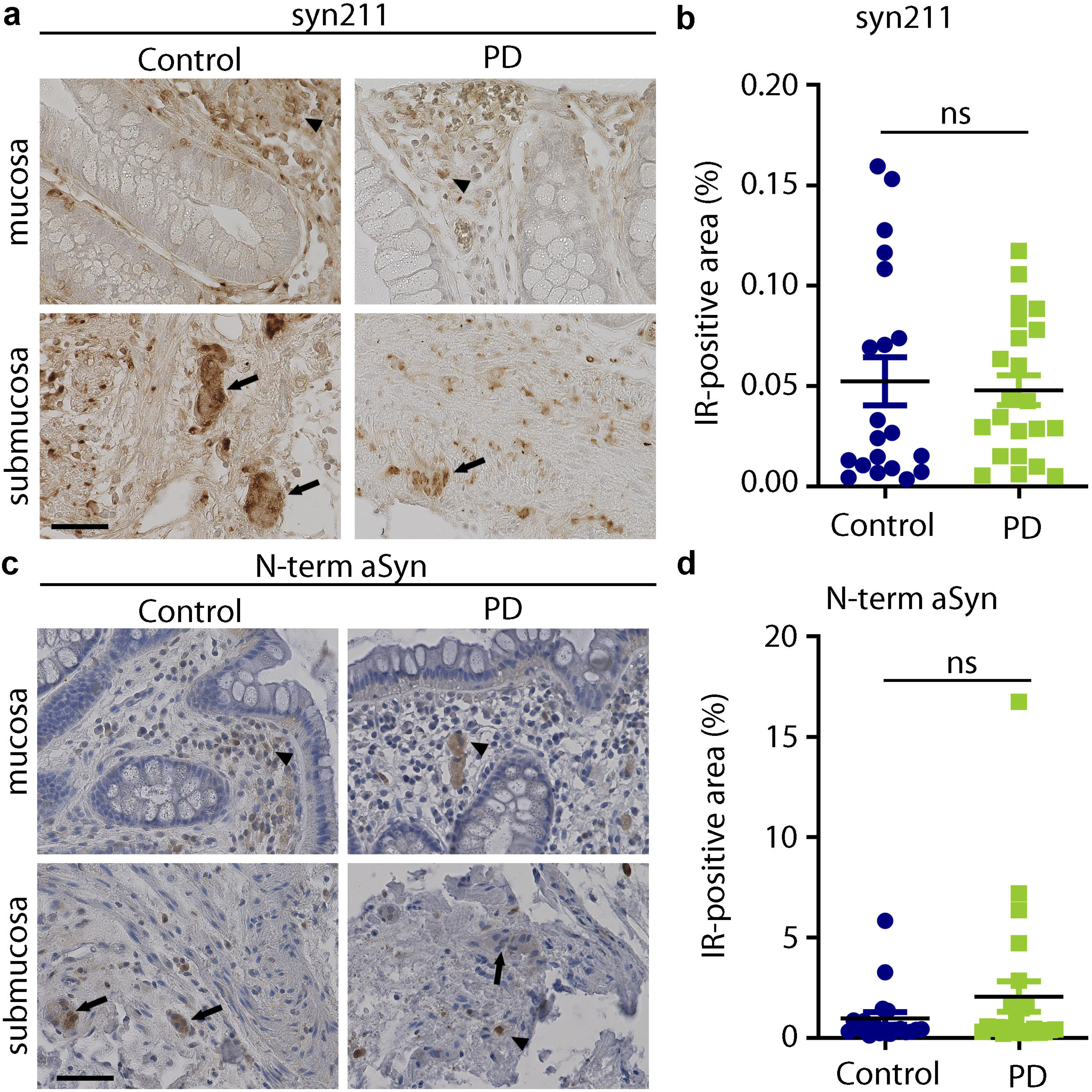
Localization pattern of aSyn in rectal biopsies of PD patients and healthy controls. Immunohistochemistry showing the distribution pattern of aSyn, clone Syn211 (a) and N-term aSyn (c) in rectal biopsies. Immunoreactivity was observed in neural structures as well as in some unidentified cells within the lamina propria. No difference was observed by morphometrical quantification of the percentage of syn211 (b) and N-term aSyn (d) immunoreactive (IR) signal to total area between PD and control (n=24 and n=20 respectively, Mann-Whitney U-test). Arrows indicate IR-positive ganglionic structures and arrowheads indicate IR-positive unidentified cells in the mucosa. Scale bar = 60 µm.

### Distribution pattern of aSyn conformers in FFPE rectal biopsies of PD patients and healthy controls

Immunoreactive signal for pathological aSyn conformers was then analysed using the MJFR-14 and 5G4 antibodies. The conformation-specific antibodies MJFR-14 and 5G4 stained for LB in the substantia nigra of the PD subjects (Fig. S4). MJFR-14 staining was not limited to LB, but also widely stained the nigral neuropil in PD patients as well as the healthy control (Fig. S4). In intestinal biopsies, MJFR-14 immunoreactivity was detected in nerve fibers and ganglia in 22 of 24 PD patients, as well as 20 of 20 of control subjects (Fig. 3a, Table 1). In most cases, MJFR-14 staining was also seen in unidentified cells in the lamina propria, most likely corresponding to immune cell populations (Fig. 3a, Table 4). The area covered by the MJFR-14 staining was not significantly changed in biopsies of PD patients in comparison to controls (Fig. 3b).

**Figure 3:**
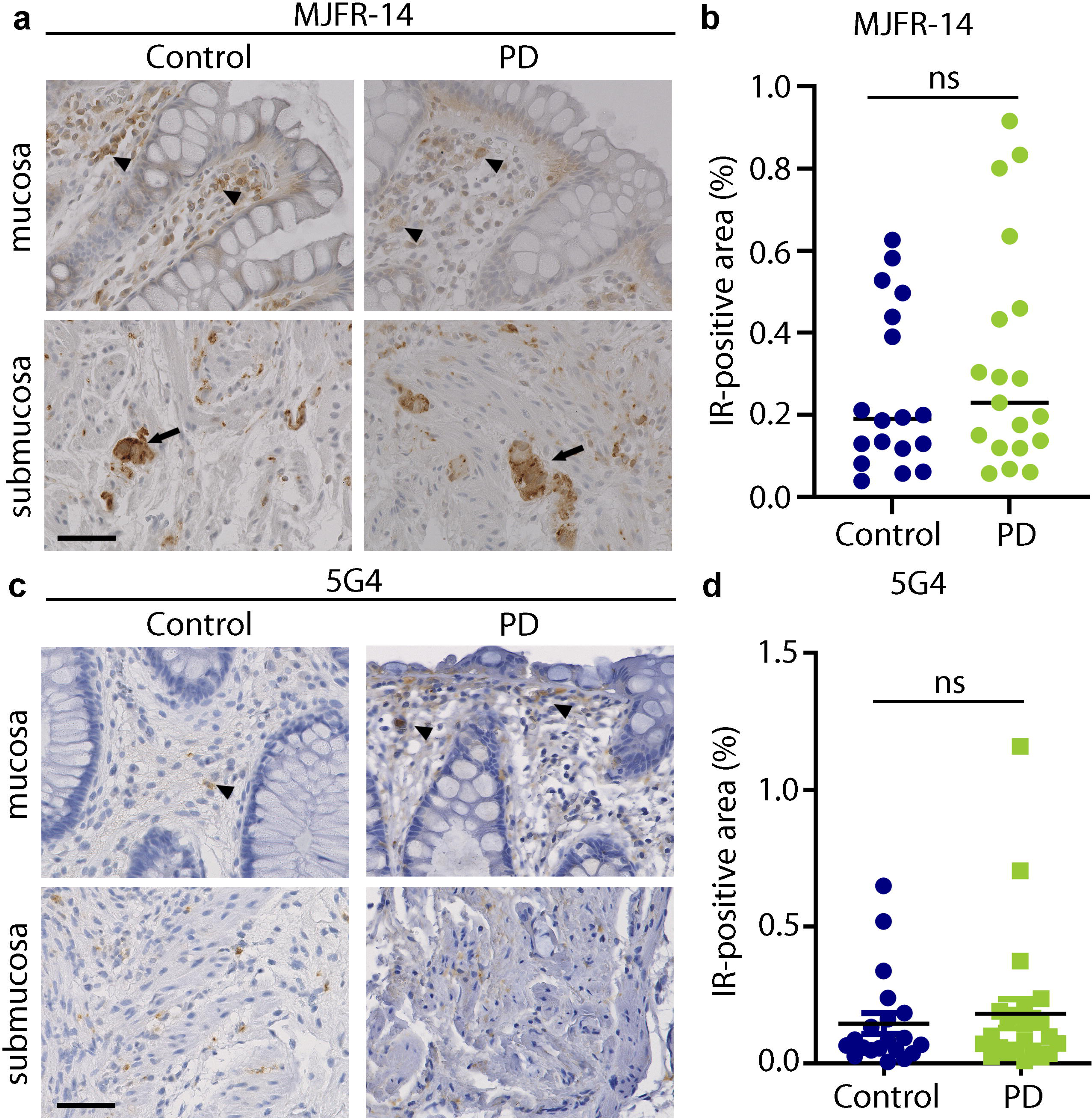
Localization pattern of conformation-specific aSyn in rectal biopsies of PD patients and healthy controls. Immunohistochemistry showing the distribution pattern of aSyn using the conformation-specific antibodies MJFR-14 (a) and clone 5G4 (c) in rectal biopsies. No difference was observed by morphometrical quantification of the percentage of MJFR-14 IR-positive signal (b) or 5G4 IR-signal (d) between PD and control (n=24 and n=20 respectively, Mann-Whitney U-test). Arrows indicate IR-positive ganglionic structures and arrowheads indicate IR-positive unidentified cells in the mucosa. Scale bar = 60 µm.

The immunoreactive signal for the 5G4 antibody was seen as disperse spots in the mucosa and submucosa in 12 of 24 PD patients and 15 of 20 healthy controls (Fig. 3c, Table 1). The staining pattern obtained for the 5G4 antibody was mainly observed in cells within the lamina propria, which may correspond to intestinal immune cells. The area covered by the 5G4 staining was not significantly altered in biopsies of PD patients in comparison to controls (Fig. 3d).

### Distribution pattern of phosphorylated-aSyn in FFPE rectal biopsies of PD patients and healthy controls

We evaluated the immunoreactive patterns of three different antibodies (EP1536Y, MJF-R13 and pSyn#64, Fig. S1) directed against phosphorylated aSyn at position S129 (P-S129 aSyn). All tested P-S129 aSyn antibodies detected LB and LN in the SN of PD patients (Fig. S4). The immunoreactive pattern of P-S129 aSyn was then assessed in FFPE rectal biopsies of PD patients and healthy controls. Whereas the signal intensity of EP1536Y aSyn remained below the limit of detection in all analyzed biopsies, both MJF-R13 and pSyn#64 clearly stained neuronal ganglia and nerve fibers within mucosal and submucosal tissue of PD and control individuals (Fig. 4, Table 1). Area covered by MJF-R13 or pSyn#64 did not differ significantly between PD biopsies and controls (Fig. 4e, f). Nonetheless, neuronal pSyn#64 immunoreactivity was significantly more present in PD patients than in the control group (75% vs. 35%, p=0.001). Noteworthy, pSyn#64 staining was correlated to Nterm-aSyn, as well as to the MJFR14 and 5G4 staining, but not to clinical parameters (Fig. S2).

**Figure 4:**
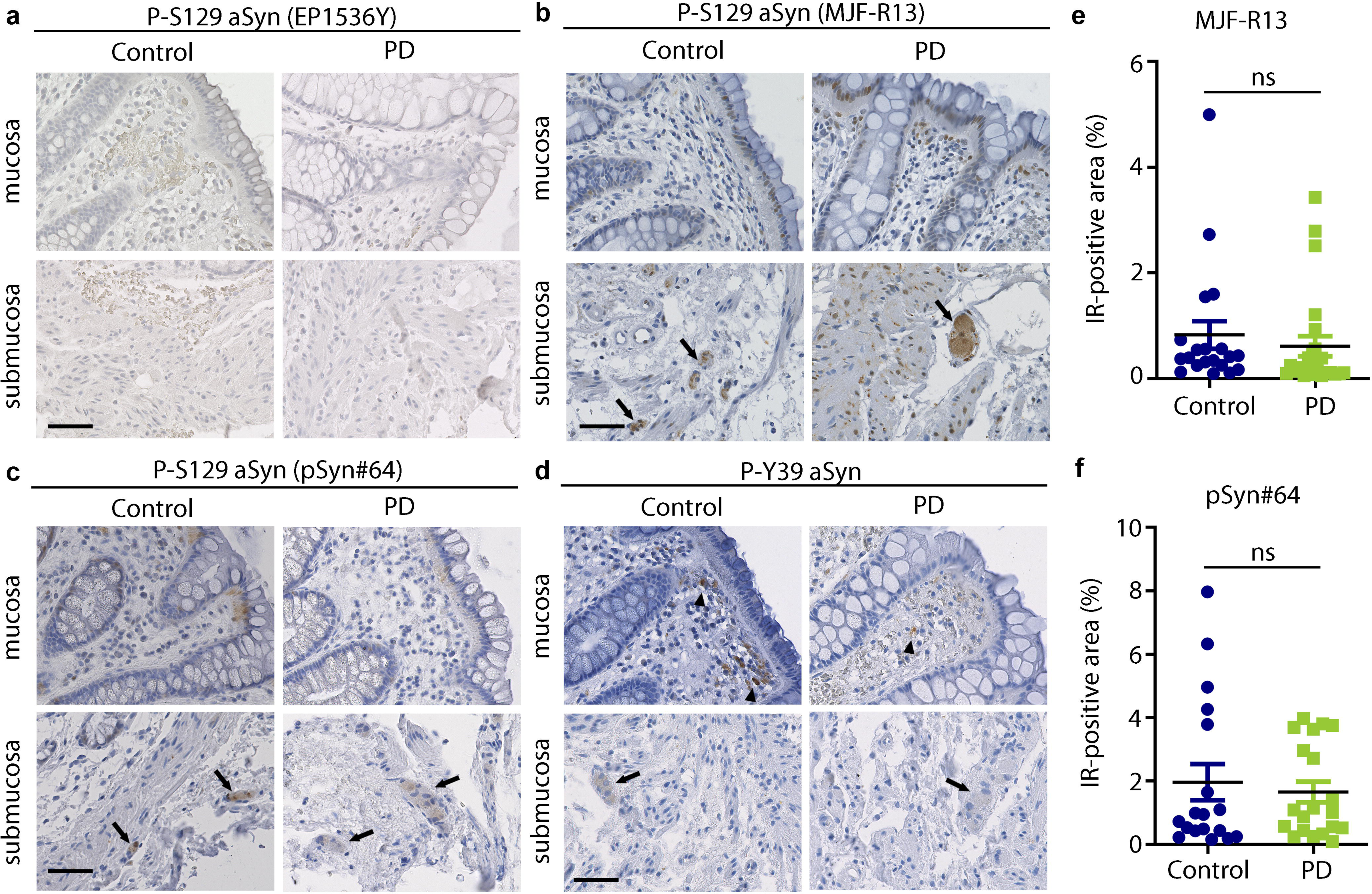
Localization pattern of phosphorylated aSyn in rectal biopsies of PD patients and healthy controls. Immunohistochemistry showing the distribution pattern of phosphorylated aSyn using the EP1536Y (a), MJF-R13 (b) and pSyn#64 (c) antibodies. No difference was observed by morphometrical quantification of the percentage of MJF-R13 IR-positive signal (e) or pSyn#64 IR-signal (f) between PD and control (n=24 and 20 respectively, Mann-Whitney U-test). Localization of P-Y39 aSyn in rectal biopsies is additionally shown (d). Arrows indicate IR-positive ganglionic structures and arrowheads indicate IR-positive unidentified cells in the mucosa. Scale bar = 60 µm.

Finally, distribution pattern of phosphorylated aSyn at position Y39 (P-Y39 aSyn) was similarly assessed. A faint immunoreactive signal for phosphorylated Y39 was observed in unidentified mucosal cell populations in the lamina propria in 18 of 20 control individuals and 19 of 24 PD patients (Fig. 4d, Table 1).

## Discussion

Considerable efforts have been made to assess the diagnostic value of misfolded aSyn in the GI tissue of PD patients. Our data indicate that tissue-specific PD-associated aSyn strains are present in rectal tissues of PD patients as demonstrated by seeding activity in an aSyn-SAA of the FFPE rectal biopsies of the PD cohort. In contrast, immunohistochemistry based on conformation- and phosphorylation-specific aSyn antibodies failed to differentiate between PD and control FFPE rectal tissues.

Earlier studies noted physiological aSyn expression in the enteric nervous system as well as in colorectal tissue (2,3,24). However, the identification of pathological aSyn deposits in these tissues has posed a greater challenge. Recent studies using alternative SAA protocols have successfully detected misfolded aSyn in intestinal tissues including duodenal and colon biopsies as well as post-mortem stomach and colon specimen of PD individuals, although with a limited sensitivity (20–22,25). In our study, SAA identified misfolded aSyn with high sensitivity *ex vivo* in the rectal biopsies of all analyzed PD patients but none of the controls. Although these data need to be confirmed in larger independent studies, they suggest that pathological aSyn strains can be detected with a high sensitivity in the rectal tissue of PD patients using an aSyn-SAA. The sensitivity of this rectal biopsy study is thus comparable to recent findings of a duodenal biopsy study and contrasts with previous debates that aSyn pathology in the GI tract of PD patients follows a rostro-caudal distribution (21,22,26). More widespread pathology in the GI tract could be explained by longer disease duration and further disease progression. However, the study presented here included individuals with a wide range of disease duration from 1 to 21 years. Since rectal biopsies can be obtained quickly and with little stress for the patient, further studies are needed to validate this finding in a larger cohort, in comparison to other biopsy sites and to different SAA methodologies. Furthermore, this study found to our knowledge for the first time a potential quantitative value of the performed intestinal aSyn-SAA, showing a positive correlation of quantitative SAA parameters with motor symptom severity (MDS-UPDRS-III scores, F_max_ and AUC) and disease duration (F_max_), which could not be found in previous studies of duodenal biopsies (22). Although these results should be interpreted with caution as a proof-of-concept, this finding suggests potential of the aSyn SAA as a progression marker, which should be confirmed in longitudinal studies.

Despite the presence of pathological aSyn as determined by SAA, we failed to detect significant pathological aSyn deposits at the histological level using established staining protocols, allowing detection of Lewy bodies and Lewy neurites in PD nigral tissues. Notably, no standard histological protocol has been developed yet to characterize pathological aSyn deposits in intestinal tissue, despite significant efforts of a dedicated consortium (12,27).

In previous studies, the conformation-specific 5G4 antibody yielded promising results in intestinal tissue. Indeed, this antibody has been used successfully to detect aSyn deposits in the colon and duodenal mucosa, yielding a sensitivity 83.3% and a specificity 88.2% in the colon, as well as a sensitivity of 100% and a specificity of 83.3% in the duodenum (15,16). In contrast to these data, we were not able to distinguish between PD patients and controls on the basis of 5G4 or MJFR-14 immunoreactivity in rectal tissue. Recent studies have indicated that the MJFR-14 antibody preferentially binds to human, patient-derived, misfolded aSyn in ELISA (17,28) as well as in dot blot assays, but loses its specificity on denatured proteins (29). Therefore, aSyn conformation may be altered in FFPE rectal biopsies and thus reduces the specificity of this antibody.

Our group and others noted the presence of P-S129 aSyn in rectal tissues of both PD patients and healthy controls in previous works (2,30). In this study, we analyzed the distribution of P-S129 aSyn using different antibodies whose specificity has been characterized in detail (31). In line with previous data, neuronal immunoreactivity observed for pSyn#64 showed the best ability to distinguish between PD and control tissue (32). In contrast, the use of the EP1536Y antibody barely demonstrated P-S129 aSyn immunoreactivity in rectal mucosal and submucosal tissues of all PD patients and controls confirming a previous study (33). It is noteworthy that immunoreactivity was observed for all patients and controls using the MJF-R13 antibodies. As mentioned above, it has been proposed that intestinal P-S129 aSyn deposits may follow a rostro-caudal gradient along the GI tract of PD patients, with the lowest values being observed in the rectum (10,26,33). Nonetheless, characterization of P-S129 aSyn in GI tissue has led to conflicting results (2,11,31,34–36). Notably, the histological analysis of P-S129 aSyn is hindered by the limited ability of existing antibodies to detect the full diversity of phosphorylated aSyn forms (31,37). Moreover, the specificity of P-S129 aSyn antibodies was shown to be dependent on aSyn truncations and additional post-translational modifications (31), which are likely to occur in the ENS (38). Similarly, significance of P-Y39 aSyn remains unclear (39–41) and the faint rectal P-Y39 aSyn staining pattern was insufficient to distinguish between PD patients and healthy controls.

It is likely that pathological aSyn deposits may be scarcely distributed within rectal tissue, as proposed elsewhere (42,43). As the ENS is an especially wide network, we may have failed to detect these sparse pathological aSyn deposits in rectal tissue during the analysis of histological sections. As proposed by Beach and co-authors, enteric neurons may be also less prone to developing aSyn pathology in the rectal ENS than in the more proximal part of the gut (10). Alternatively, the positive histological aSyn signal observed in the proximal GI tract in previous studies may correspond to extrinsic vagal nerve fibers, which are likely absent in rectal tissue (7,10). Differences in tissue location (proximal vs. distal GI tract) and processing (fresh vs. FFPE biopsies), staining protocols and antibodies may further explain the discrepancies observed between different studies. Finally, it has to be acknowledged that the gender distribution between the control and PD group differed significantly in the presented study. Although it cannot be assumed that gender has any influence on the presented results, further studies should also strive for a matching in this respect.

Taken together, it is tempting to speculate that the pathological aSyn conformation in rectal tissue may differ from nigral aSyn at the biochemical level and that predominantly early oligomeric aSyn strains are present and detected by SAA in rectal biopsies. This assumption is in line with recent models of PD subtyping, supporting the presence of tissue-specific pathological aSyn strains (44–47). Our data emphasize the need for further characterization of the biochemical conformation of aSyn strains, including aSyn post-translational modifications in both the enteric/autonomic and the central nervous system (31,48,49).

In conclusion, our data suggest that SAA is better suited to detect the full spectrum of pathological aSyn conformers in rectal tissue than histological approaches. SAA and the development of aSyn strain-specific detection methods may provide further crucial insights into PD pathology along the gut-brain axis. Moreover, future studies should investigate changes of the quantitative a-Syn-SAA parameters derived from GI tissue during the course of the disease.

## Data Availability

All data produced in the present study are available upon reasonable request to the authors

## Acknowledgment

The authors thank Katrin Neblung-Masuhr, Lisa Behnke, Fabian Neumann and Dagmar Lukas (Institute for Anatomy, Kiel University) for their excellent technical assistance. The authors are grateful to Dr. Neele Schumacher (Biochemical Institute, Kiel University) for providing access to and technical support in the use of the Leica slide scanner. This project was supported by a grant of the Faculty of Medicine, Kiel University (K126422) to FC and ES. Funders and supporting institutions had no influence on the design, conduct, or analysis of the study. We thank all patients for their participation in this study.

## Relevant conflicts of interest/financial disclosures

The authors declare no conflict of interest. Funders and supporting institutions had no influence on the design, conduct, or analysis of the study.

## Authors’ Contributions

Annika Kluge: study concept and design, acquisition and interpretation of SAA data, statistical analysis, drafting of the manuscript. Carmen Kintrup, Katja Schröder: acquisition and interpretation of histology data, statistical analysis. Kristina Kulcsarova, Julius Welzel, Sebastian Heinzel: interpretation of data, statistical analysis, critical revision of the manuscript for intellectual content. Sarah K Bonkat, Manuela Pendziwiat, Stephan Schoch: acquisition of data, study supervision. Martina Böttner, Mark Ellrichmann: acquisition of data, study supervision, critical revision of the manuscript for intellectual content. Thilo Wedel, Ralph Lucius, Daniela Berg: study supervision, critical revision of the manuscript for intellectual content. Eva Schaeffer: study concept and design, study supervision, acquisition and interpretation of data, critical revision of the manuscript for intellectual content. François Cossais: study concept and design, study supervision, acquisition and interpretation of histology data, statistical analysis, drafting of the manuscript.

## Notes

### Competing Interest Statement

The authors have declared no competing interest.

### Author Declarations

The study was approved by the local ethics committee of the faculty of medicine, Christian-Albrechts University of Kiel, Germany (D452/19).

### Summary of Updates

Histology data of the evaluation of phosphorylated alpha-synuclein immunoreactivity have been updated and now summarize in table 1. Proximity ligation assay analysis has been withdrawed of this version.

## References

1. Ivan I-F, Irincu V-L, Diaconu Ștefania, Falup-Pecurariu O, Ciopleiaș B, Falup-Pecurariu C. Gastro-intestinal dysfunctions in Parkinson’s disease (Review). Exp Ther Med [Internet]. 2021 Oct;22(4):1083. Available from: http://www.ncbi.nlm.nih.gov/pubmed/34447476

2. Barrenschee M, Zorenkov D, Böttner M, Lange C, Cossais F, Scharf AB, et al. Distinct pattern of enteric phospho-alpha-synuclein aggregates and gene expression profiles in patients with Parkinson’s disease. Acta Neuropathol Commun [Internet]. 2017 Dec 5;5(1):1. Available from: 10.1186/s40478-016-0408-2

3. Böttner M, Zorenkov D, Hellwig I, Barrenschee M, Harde J, Fricke T, et al. Expression pattern and localization of alpha-synuclein in the human enteric nervous system. Neurobiol Dis [Internet]. 2012 Dec [cited 2012 Nov 11];48(3):474–80. Available from: http://www.ncbi.nlm.nih.gov/pubmed/22850485

4. Cossais F, Schaeffer E, Heinzel S, Zimmermann J, Niesler B, Röth R, et al. Expression Profiling of Rectal Biopsies Suggests Altered Enteric Neuropathological Traits in Parkinson’s Disease Patients. J Parkinsons Dis [Internet]. 2021 Dec 16;11(1):171–6. Available from: https://www.medra.org/servlet/aliasResolver?alias=iospress&doi=10.3233/JPD-202258

5. Schaeffer E, Kluge A, Böttner M, Zunke F, Cossais F, Berg D, et al. Alpha Synuclein Connects the Gut-Brain Axis in Parkinson’s Disease Patients - A View on Clinical Aspects, Cellular Pathology and Analytical Methodology. Front cell Dev Biol [Internet]. 2020 Sep 8;8:573696. Available from: https://www.frontiersin.org/article/10.3389/fcell.2020.573696/full

6. Derkinderen P, Cossais F, de Guilhem de Lataillade A, Leclair-Visonneau L, Neunlist M, Paillusson S, et al. Gastrointestinal mucosal biopsies in Parkinson’s disease: beyond alpha-synuclein detection. J Neural Transm [Internet]. 2022 Sep 24;129(9):1095–103. Available from: http://www.ncbi.nlm.nih.gov/pubmed/34816335

7. Braak H, Rüb U, Gai WP, Del Tredici K. Idiopathic Parkinson’s disease: possible routes by which vulnerable neuronal types may be subject to neuroinvasion by an unknown pathogen. J Neural Transm [Internet]. 2003 May [cited 2014 Oct 7];110(5):517–36. Available from: http://www.ncbi.nlm.nih.gov/pubmed/12721813

8. Schneider SA, Boettner M, Alexoudi A, Zorenkov D, Deuschl G, Wedel T. Can we use peripheral tissue biopsies to diagnose Parkinson’s disease? A review of the literature. Eur J Neurol [Internet]. 2016 Feb;23(2):247–61. Available from: http://www.ncbi.nlm.nih.gov/pubmed/26100920

9. Bu J, Liu J, Liu K, Wang Z. Diagnostic utility of gut α-synuclein in Parkinson’s disease: A systematic review and meta-analysis. Behav Brain Res [Internet]. 2019 May;364:340–7. Available from: https://linkinghub.elsevier.com/retrieve/pii/S0166432818312063

10. Beach TG, Adler CH, Sue LI, Vedders L, Lue L, White Iii CL, et al. Multi-organ distribution of phosphorylated alpha-synuclein histopathology in subjects with Lewy body disorders. Acta Neuropathol [Internet]. 2010 Jun [cited 2014 Aug 19];119(6):689–702. Available from: http://www.pubmedcentral.nih.gov/articlerender.fcgi?artid=2866090&tool=pmcentrez&rendertype=abstract

11. Ruffmann C, Bengoa-Vergniory N, Poggiolini I, Ritchie D, Hu MT, Alegre-Abarrategui J, et al. Detection of alpha-synuclein conformational variants from gastro-intestinal biopsy tissue as a potential biomarker for Parkinson’s disease. Neuropathol Appl Neurobiol. 2018;44(7):722–36.

12. Chahine LM, Beach TG, Brumm MC, Adler CH, Coffey CS, Mosovsky S, et al. In vivo distribution of α-synuclein in multiple tissues and biofluids in Parkinson disease. Neurology [Internet]. 2020 Sep 1;95(9):e1267–84. Available from: http://www.ncbi.nlm.nih.gov/pubmed/32747521

13. Fujiwara H, Hasegawa M, Dohmae N, Kawashima A, Masliah E, Goldberg MS, et al. alpha-Synuclein is phosphorylated in synucleinopathy lesions. Nat Cell Biol [Internet]. 2002 Feb;4(2):160–4. Available from: http://www.ncbi.nlm.nih.gov/pubmed/11813001

14. Kovacs GG, Wagner U, Dumont B, Pikkarainen M, Osman AA, Streichenberger N, et al. An antibody with high reactivity for disease-associated α-synuclein reveals extensive brain pathology. Acta Neuropathol. 2012;124(1):37–50.

15. Emmi A, Sandre M, Russo FP, Tombesi G, Garrì F, Campagnolo M, et al. Duodenal alpha-Synuclein Pathology and Enteric Gliosis in Advanced Parkinson’s Disease. Mov Disord. 2023 May;38(5):885–94.

16. Skorvanek M, Gelpi E, Mechirova E, Ladomirjakova Z, Han V, Lesko N, et al. α-Synuclein antibody 5G4 identifies manifest and prodromal Parkinson’s disease in colonic mucosa. Mov Disord [Internet]. 2018;33(8):1366–8. Available from: http://www.ncbi.nlm.nih.gov/pubmed/30230627

17. Kumar ST, Jagannath S, Francois C, Vanderstichele H, Stoops E, Lashuel HA. How specific are the conformation-specific α-synuclein antibodies? Characterization and validation of 16 α-synuclein conformation-specific antibodies using well-characterized preparations of α-synuclein monomers, fibrils and oligomers with distinct struct. Neurobiol Dis [Internet]. 2020;146(August):105086. Available from: 10.1016/j.nbd.2020.105086

18. Saijo E, Groveman BR, Kraus A, Metrick M, Orrù CD, Hughson AG, et al. Ultrasensitive RT-QuIC Seed Amplification Assays for Disease-Associated Tau, α-Synuclein, and Prion Aggregates. Methods Mol Biol [Internet]. 2019;1873:19–37. Available from: http://www.ncbi.nlm.nih.gov/pubmed/30341601

19. Roostaee A, Beaudoin S, Staskevicius A, Roucou X. Aggregation and neurotoxicity of recombinant α-synuclein aggregates initiated by dimerization. Mol Neurodegener [Internet]. 2013 Jan 22;8:5. Available from: http://www.ncbi.nlm.nih.gov/pubmed/23339399

20. Bargar C, Wang W, Gunzler SA, LeFevre A, Wang Z, Lerner AJ, et al. Streamlined alpha-synuclein RT-QuIC assay for various biospecimens in Parkinson’s disease and dementia with Lewy bodies. Acta Neuropathol Commun [Internet]. 2021;9(1):1–13. Available from: 10.1186/s40478-021-01175-w

21. Fenyi A, Leclair-Visonneau L, Clairembault T, Coron E, Neunlist M, Melki R, et al. Detection of alpha-synuclein aggregates in gastrointestinal biopsies by protein misfolding cyclic amplification. Neurobiol Dis [Internet]. 2019 Sep;129(April):38–43. Available from: 10.1016/j.nbd.2019.05.002

22. Vascellari S, Orrù CD, Groveman BR, Parveen S, Fenu G, Pisano G, et al. α-Synuclein seeding activity in duodenum biopsies from Parkinson’s disease patients. PLOS Pathog. 2023;19(6):e1011456.

23. Postuma RB, Berg D, Stern M, Poewe W, Olanow CW, Oertel W, et al. MDS clinical diagnostic criteria for Parkinson’s disease. Mov Disord [Internet]. 2015 Oct 16 [cited 2015 Oct 19];30(12):1591–601. Available from: http://www.ncbi.nlm.nih.gov/pubmed/26474316

24. Antunes L, Frasquilho S, Ostaszewski M, Weber J, Longhino L, Antony P, et al. Similar α-Synuclein staining in the colon mucosa in patients with Parkinson’s disease and controls. Mov Disord [Internet]. 2016;31(10):1567–70. Available from: http://www.ncbi.nlm.nih.gov/pubmed/27324838

25. Fenyi A, Duyckaerts C, Bousset L, Braak H, Del Tredici K, Melki R. Seeding propensity and characteristics of pathogenic αsyn assemblies in formalin-fixed human tissue from the enteric nervous system, olfactory bulb, and brainstem in cases staged for parkinson’s disease. Cells. 2021;10(1):1–20.

26. Pouclet H, Lebouvier T, Coron E, des Varannes SB, Rouaud T, Roy M, et al. A comparison between rectal and colonic biopsies to detect Lewy pathology in Parkinson’s disease. Neurobiol Dis [Internet]. 2012 Jan [cited 2014 Oct 8];45(1):305–9. Available from: http://www.ncbi.nlm.nih.gov/pubmed/21878391

27. Corbillé A-GG, Letournel F, Kordower JH, Lee J, Shanes E, Neunlist M, et al. Evaluation of alpha-synuclein immunohistochemical methods for the detection of Lewy-type synucleinopathy in gastrointestinal biopsies. Acta Neuropathol Commun [Internet]. 2016 Apr 4;4:35. Available from: http://www.ncbi.nlm.nih.gov/pubmed/27044604

28. Berkhoudt Lassen L, Gregersen E, Kathrine Isager A, Betzer C, Hahn Kofoed R, Henning Jensen P. ELISA method to detect α-synuclein oligomers in cell and animal models. PLoS One. 2018;13(4):1–12.

29. Kluge A, Bunk J, Schaeffer E, Drobny A, Xiang W, Knacke H, et al. Detection of neuron-derived pathological α-synuclein in blood. Brain [Internet]. 2022;145(9):3058–71. Available from: http://www.ncbi.nlm.nih.gov/pubmed/35722765

30. Beach TG, Serrano GE, Kremer T, Canamero M, Dziadek S, Sade H, et al. Immunohistochemical Method and Histopathology Judging for the Systemic Synuclein Sampling Study (S4). J Neuropathol Exp Neurol [Internet]. 2018 Sep 1;77(9):793–802. Available from: https://academic.oup.com/jnen/article/77/9/793/5063596

31. Lashuel HA, Mahul-Mellier AL, Novello S, Hegde RN, Jasiqi Y, Altay MF, et al. Revisiting the specificity and ability of phospho-S129 antibodies to capture alpha-synuclein biochemical and pathological diversity. npj Park Dis. 2022;8(1):1–19.

32. Sadoc M, Clairembault T, Coron E, Berthomier C, Le Dily S, Vavasseur F, et al. Wake and non-rapid eye movement sleep dysfunction is associated with colonic neuropathology in Parkinson’s disease. Sleep [Internet]. 2024 Mar 11;47(3). Available from: http://www.ncbi.nlm.nih.gov/pubmed/38156524

33. Shin C, Park S-H, Yun JY, Shin JH, Yang H-K, Lee H-J, et al. Fundamental limit of alpha-synuclein pathology in gastrointestinal biopsy as a pathologic biomarker of Parkinson’s disease: Comparison with surgical specimens. Parkinsonism Relat Disord [Internet]. 2017 Nov;44:73–8. Available from: http://www.ncbi.nlm.nih.gov/pubmed/28923294

34. Lebouvier T, Neunlist M, Bruley des Varannes S, Coron E, Drouard A, N’Guyen J-M, et al. Colonic biopsies to assess the neuropathology of Parkinson’s disease and its relationship with symptoms. PLoS One [Internet]. 2010 Jan [cited 2012 Nov 11];5(9):e12728. Available from: http://www.pubmedcentral.nih.gov/articlerender.fcgi?artid=2939055&tool=pmcentrez&rendertype=abstract

35. Lebouvier T, Chaumette T, Damier P, Coron E, Touchefeu Y, Vrignaud S, et al. Pathological lesions in colonic biopsies during Parkinson’s disease. Gut [Internet]. 2008 Dec [cited 2014 Oct 8];57(12):1741–3. Available from: http://www.ncbi.nlm.nih.gov/pubmed/19022934

36. Visanji NP, Marras C, Kern DS, Al Dakheel A, Gao A, Liu LWC, et al. Colonic mucosal-synuclein lacks specificity as a biomarker for Parkinson disease. Neurology [Internet]. 2015 Feb 10;84(6):609–16. Available from: http://www.neurology.org/cgi/doi/10.1212/WNL.0000000000001240

37. Delic V, Chandra S, Abdelmotilib H, Maltbie T, Wang S, Kem D, et al. Sensitivity and specificity of phospho-Ser129 α-synuclein monoclonal antibodies. J Comp Neurol [Internet]. 2018 Aug 15;526(12):1978–90. Available from: http://www.ncbi.nlm.nih.gov/pubmed/29888794

38. Killinger BA, Madaj Z, Sikora JW, Rey N, Haas AJ, Vepa Y, et al. The vermiform appendix impacts the risk of developing Parkinson’s disease. Sci Transl Med [Internet]. 2018 Oct 31;10(465). Available from: http://www.ncbi.nlm.nih.gov/pubmed/30381408

39. Mahul-Mellier AL, Fauvet B, Gysbers A, Dikiy I, Oueslati A, Georgeon S, et al. C-Abl phosphorylates α-synuclein and regulates its degradation: Implication for α-synuclein clearance and contribution to the pathogenesis of parkinson’s disease. Hum Mol Genet. 2014;23(11):2858–79.

40. Brahmachari S, Ge P, Lee SH, Kim D, Karuppagounder SS, Kumar M, et al. Activation of tyrosine kinase c-Abl contributes to α-synuclein-induced neurodegeneration. J Clin Invest. 2016;126(8):2970–88.

41. Altay MF, Liu AKL, Holton JL, Parkkinen L, Lashuel HA. Prominent astrocytic alpha-synuclein pathology with unique post-translational modification signatures unveiled across Lewy body disorders. Acta Neuropathol Commun. 2022;10(1):1–18.

42. Borghammer P. The α-Synuclein Origin and Connectome Model (SOC Model) of Parkinson’s Disease: Explaining Motor Asymmetry, Non-Motor Phenotypes, and Cognitive Decline. J Parkinsons Dis [Internet]. 2021;11(2):455–74. Available from: http://www.ncbi.nlm.nih.gov/pubmed/33682732

43. Borghammer P. How does parkinson’s disease begin? Perspectives on neuroanatomical pathways, prions, and histology. Mov Disord. 2018;33(1):48–57.

44. Just MK, Gram H, Theologidis V, Jensen PH, Nilsson KPR, Lindgren M, et al. Alpha-Synuclein Strain Variability in Body-First and Brain-First Synucleinopathies. Front Aging Neurosci. 2022;14(May):1–18.

45. Corbillé AG, Neunlist M, Derkinderen P. Cross-linking for the analysis of α-synuclein in the enteric nervous system. J Neurochem. 2016;139(5):839–47.

46. Hass EW, Sorrentino ZA, Xia Y, Lloyd GM, Trojanowski JQ, Prokop S, et al. Disease-, region- and cell type specific diversity of α-synuclein carboxy terminal truncations in synucleinopathies. Acta Neuropathol Commun [Internet]. 2021 Aug 28;9(1):146. Available from: http://www.ncbi.nlm.nih.gov/pubmed/34454615

47. Chalazonitis A, Rao M, Sulzer D. Similarities and differences between nigral and enteric dopaminergic neurons unravel distinctive involvement in Parkinson’s disease. NPJ Park Dis [Internet]. 2022 Apr 22;8(1):50. Available from: http://www.ncbi.nlm.nih.gov/pubmed/35459867

48. Zhang S, Zhu R, Pan B, Xu H, Olufemi MF, Gathagan RJ, et al. Post-translational modifications of soluble α-synuclein regulate the amplification of pathological α-synuclein. Nat Neurosci [Internet]. 2023 Feb;26(2):213–25. Available from: http://www.ncbi.nlm.nih.gov/pubmed/36690898

49. Altay MF, Kumar ST, Burtscher J, Jagannath S, Strand C, Miki Y, et al. Development and validation of an expanded antibody toolset that captures alpha-synuclein pathological diversity in Lewy body diseases. bioRxiv. 2022;preprint.

