## Supplementary information for "Alpha-synuclein distribution and seeding activity in rectal biopsies in Parkinson’s disease"

Supplementary table 1. List of antibodies used in this study

| **Antibody / Clone** | **Ref.** | **Dilution** | **Lot** | **RRID** | **Supplier** |
|---|---|---|---|---|---|
| mouse anti-aSyn clone Syn211 | ab80627 | 1:1000 | GR3324402-1 | RRID:AB_1603277 | Abcam |
| rabbit anti-N-Terminal aSyn | ab6176 | 1:5000 | | RRID:AB_305344 | Abcam |
| rabbit anti-fibrillary aSyn MJFR-14-6-4-2 | ab209538 | 1:2000 | GR3256670-3 | RRID:AB_2714215 | Abcam |
| mouse anti-aSyn clone 5G4 | MABN389 | 1:500 | VP2201053 | RRID:AB_2716647 | Millipore |
| rabbit anti-P-S129 aSyn EP1536Y | ab51253 | 1:200 | GR3437967-8 | RRID:AB_869973 | Abcam |
| rabbit anti-P-S129 aSyn MJF-R13 | ab168381 | 1:500 | 1015267-2 | RRID:AB_2728613 | Abcam |
| mouse anti-P-S129 aSyn pSyn#64 | 014-20281 | 1:1000 | STH6983 | RRID:AB_516843 | Wako chemicals |
| mouse anti-phosphorylated Y39 aSyn | 849202 | 1:500 | B253561 | RRID:AB_2650702 | Biolegend |

Supplementary results

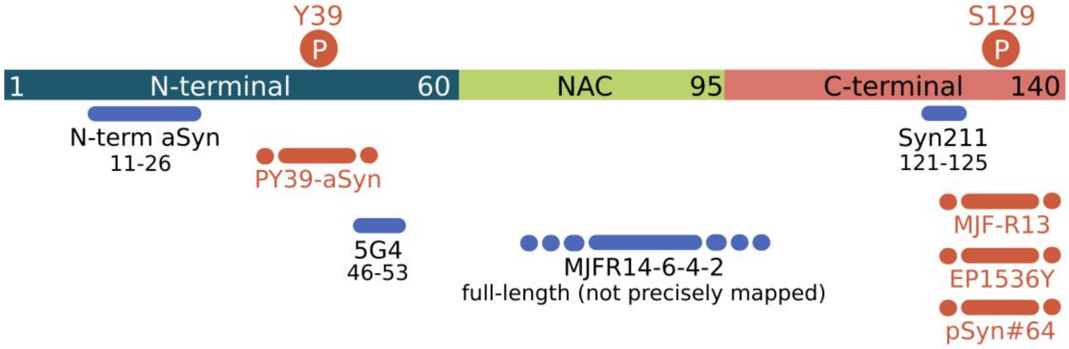


**Supp. Figure 1:** Scheme representing known binding sites of aSyn antibodies used in this study on the aSyn protein sequence.

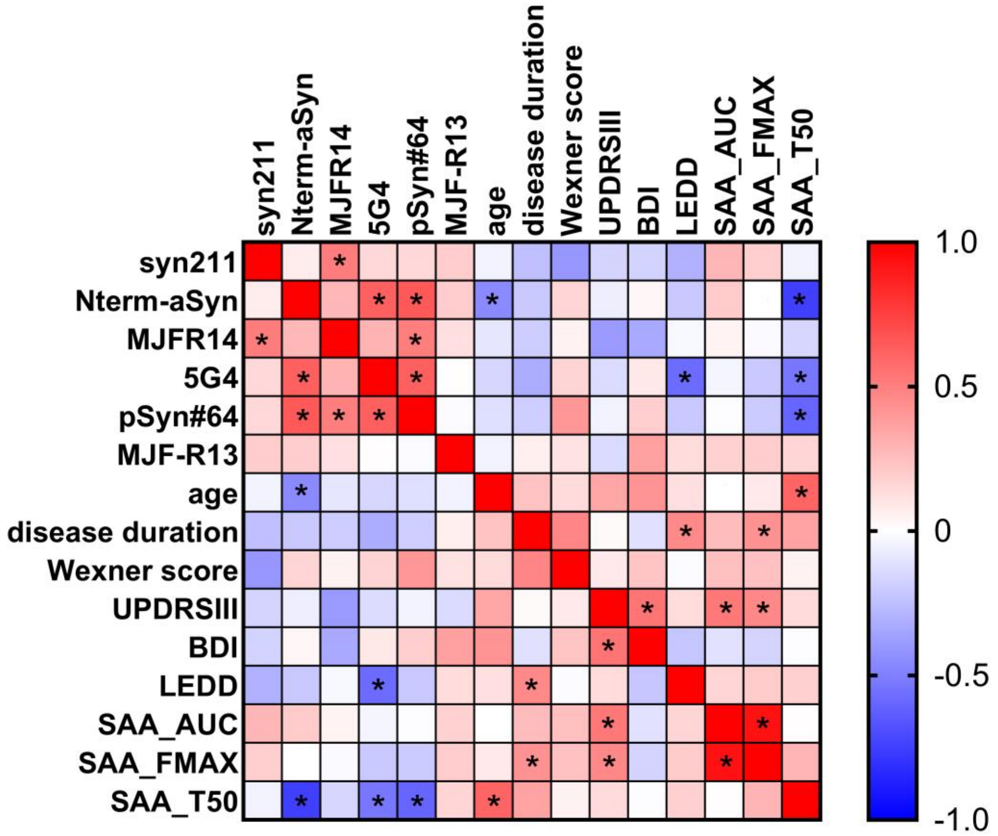


**Supp. Figure 2:** Correlation matrix between clinical parameters, immunohistochemistry and SAA parameters (AUC, Fmax and T50) in the Parkinson's disease group. Asterisk indicates a statistically significant correlation (Spearman's correlation, $p < 0.05$).

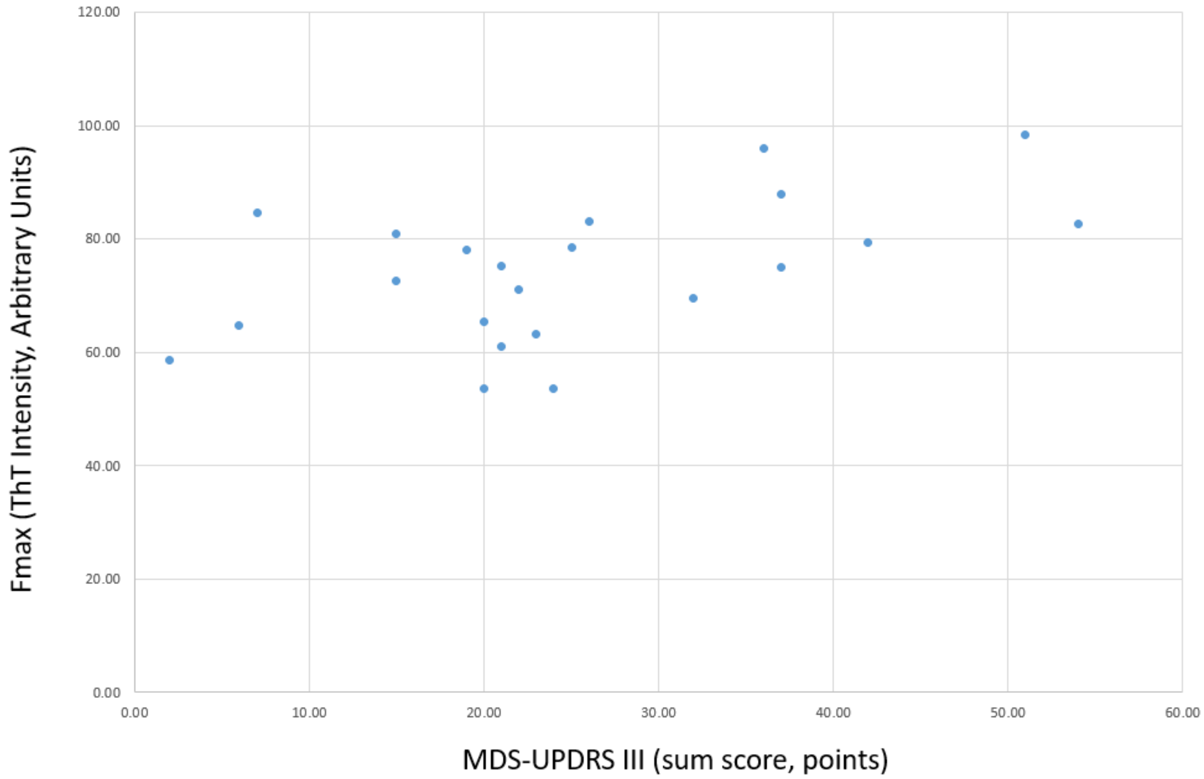


**Supp. Figure 3:** Correlation between MDS-UPDRSIII (sum score) and SAA Fmax (ThT intensity, arbitrary units) for the Parkinson's disease group. Spearman's correlation: r = 0.463, p = 0.03.

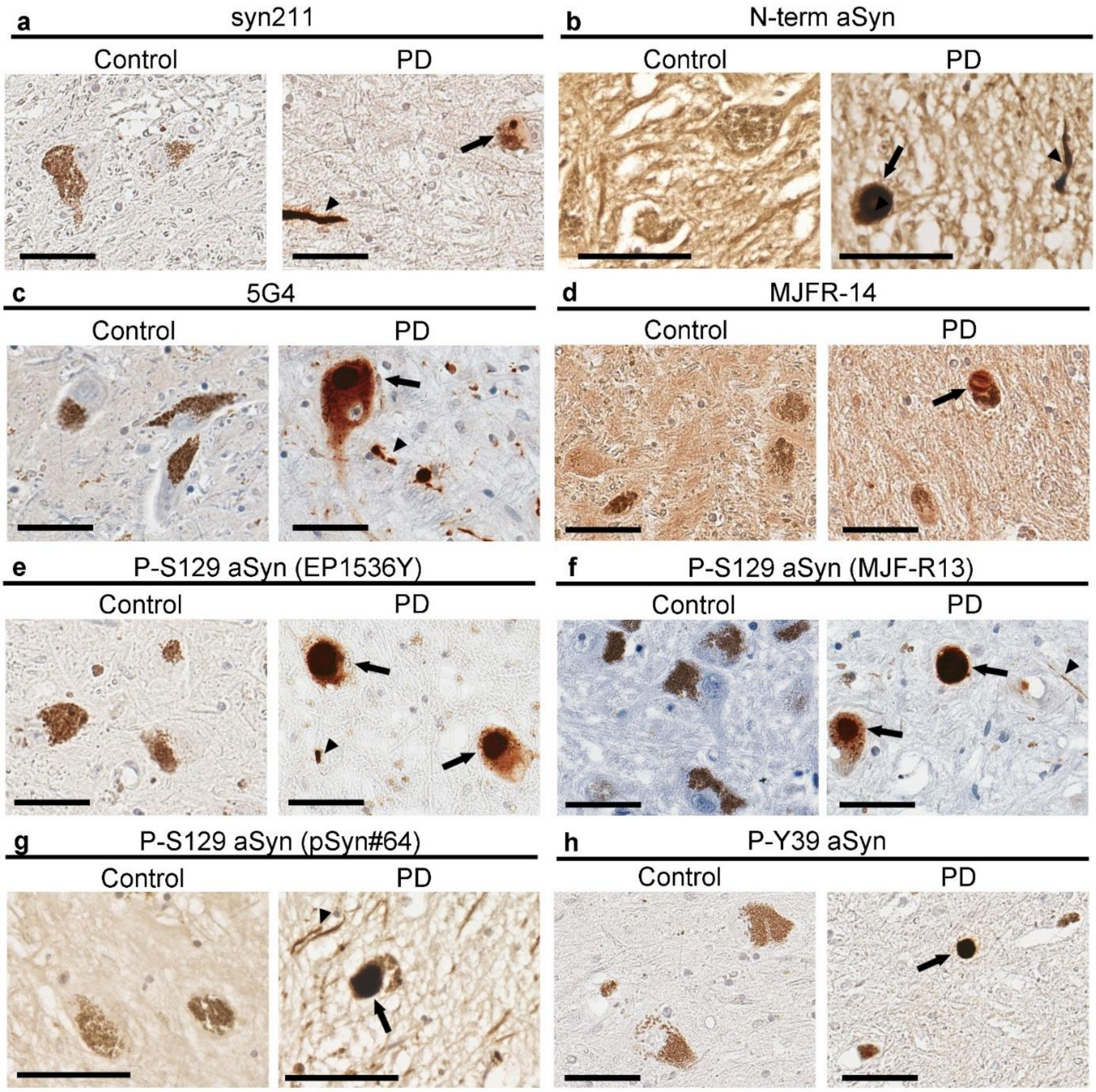


**Supp. Figure 4:** Localization pattern of a) syn211, b) N-term aSyn, c) clone 5G4, d) MJFR-14, e) EP1536Y, f) MJF-R13, g) pSyn#64 and h) P-Y39 aSyn in the human substantia nigra of one PD patient and one control individual. Immunoreactivity was observed in Lewy bodies (black arrow), as well as in Lewy neurites (black arrowheads). scale bar = 60 µm.

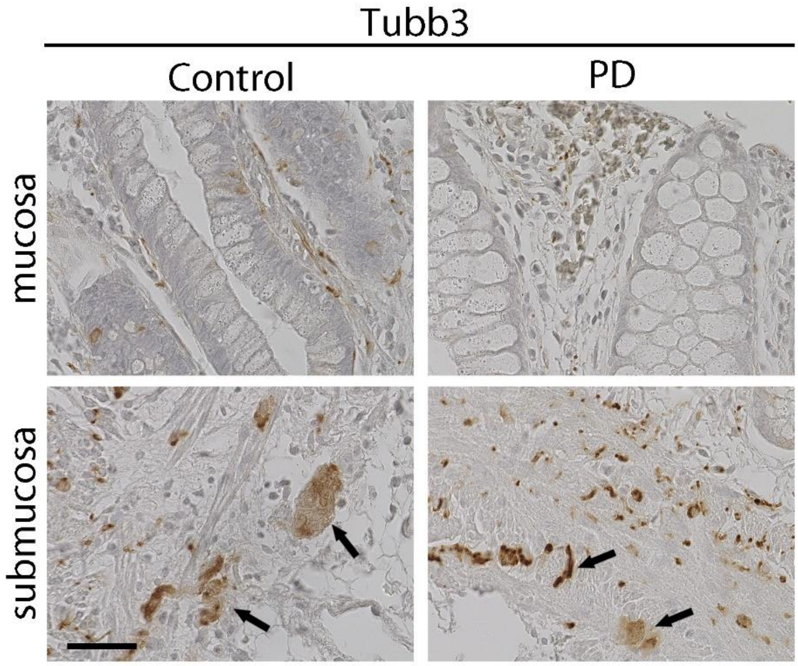


**Supp. Figure 5:** Localization of Tubb3 in rectal mucosa and submucosa of PD patients and controls. Black arrows point to immunoreactive signal in ganglionic structures of the submucosa. scale bar = 60 µm.
